# Understanding the impact of a social support program in Immokalee, FL, during the COVID-19 pandemic

**DOI:** 10.1101/2023.02.23.23286348

**Authors:** Lindsay Richards, Leping Wang, Joashilia Jeanmarie, Shirin Shafazand, Daniel Palazuelos, Vitina Monacello

## Abstract

From December 2020 to July 2021, $700,000 was distributed in direct cash transfers to residents of Immokalee, FL who tested positive for COVID-19. The goal of this study is to evaluate the impact of this cash transfer program. We conducted 157 structured interviews with program beneficiaries via phone call or home visit and asked about sociodemographic variables, how the money was used, whether the money was sufficient for two weeks’ financial needs, and participant ability to self-isolate. A logit regression model was then used to explore the relationships between sociodemographic variables and whether the respondent thought the money was enough for two weeks of financial needs. A majority of respondents (83.7%) reported spending the check exclusively on living expenses, and 99.3% reported that the money helped them stay home and quarantine while having COVID-19. Offering direct cash transfers of $800-$1200 to residents of Immokalee, FL who tested positive for COVID-19 was effective in reducing COVID-associated financial burden, and this money was most likely to be spent on living necessities rather than temptation goods. People with housing insecurity and without a high school degree were significantly less likely to report that the money was enough for two weeks’ financial needs, indicating that these characteristics mark those in the population who may have needed more support. Given that the COVID-19 pandemic has exacerbated pre-existing health disparities, it is important to understand the role of cash transfers as a public health tool and their potential impact on community mitigation efforts.

## Introduction

According to the Mixteco Indigena Community Organizing Project (MICOP) and Central Coast Alliance for a Sustainable Economy (CAUSE), “farm workers remain the disposable essential worker” [1]. From plantation economies to the present-day farm work industry, agribusiness in Florida has relied on extracting wealth from workers who are deemed expendable and excluded from rights and protections offered to the rest of the population. These fault lines can be exacerbated by disaster, as was the case when the COVID-19 pandemic arrived in Immokalee. Immokalee, FL, is an agricultural community located in Collier County of Southwest Florida known for its tomato and citrus production. The number of residents changes from approximately 14,000 during the off-season to between 20,000 and 30,000 during the tomato picking season, which runs from September to June [2]. Its population is composed largely of Mexican, Guatemalan, and Haitian immigrants, with nearly 40% of residents living below the poverty line [3]. Farmworkers face many structural inequities that make them especially vulnerable to the health and economic consequences of COVID-19 – crowded housing conditions, poor labor protections, financial precarity, and documentation status, among others [4]. By mid-June of 2020, Immokalee had reported 899 positive cases out of 2500 tests conducted, while in neighboring Naples - a wealthier zip code but within the same county - there had been only 76 cases [5]. Driven by advocacy from the Coalition of Immokalee Workers (CIW), a worker-based human rights organization, many different not-for-profit groups stepped in to respond to this crisis, including a local nonprofit called Misión Peniel. Misión Peniel’s response focused on addressing the financial burden of missing two weeks of work for quarantine purposes by implementing a cash transfer program for individuals who tested positive for COVID-19. The aim of this cash distribution was to facilitate self-isolation while COVID-19 positive, incentivize testing, and help alleviate the disproportionate impact of the virus in the Immokalee community. Residents of Immokalee were able to apply for this funding in two ways. The first is that individuals were referred directly from COVID-19 testing events and their applications were processed onsite after they received counseling regarding positive results and public health recommendations. The second way to apply was to call a hotline number that was set up for individuals who were tested elsewhere but lived in Immokalee. A team member would then return their call to assist with completing an application over the phone. The eligibility requirements for the cash transfer program were that recipients must be Immokalee residents and must provide proof of current COVID-19 infection documented by a positive test in the past 10 days (per CDC guideline at the time). Applications were tracked in a spreadsheet and were evaluated in the order in which they were entered on an ongoing basis. Applications were reviewed by Mission Peniel staff to verify proof of eligibility namely address/ID provided and documentation of positive test results. Less than 5% of applicants were denied. Reasons for being denied included falsifying information on the application, not submitting proof of address/residency in Immokalee, or lack of verification of positive test results. From December 2020 to July 2021, Misión Peniel distributed one-time checks of $800 for single applicants and $1000 or $1200 for applicants with children. The initial amount for families, defined as a parent caring for a minor child/children, was set at $1200. However, within a few weeks of the program the amount was reduced to $1000 in order to stretch limited funding during a surge in cases. A total of 778 checks were distributed during this time period. Most participants received a check within 7 days of applying, but some applications experienced additional administrative delays. Checks were distributed one to two times a week and were available for pick-up outside of Misión Peniel’s office or delivered to recipients’ homes. Given that many individuals were quarantining at the time of distribution, a family member or friend was able to pick up the check for them. Assistance was provided in the form of checks because this was the most accessible means of receiving funds for the community.

## Methods

### Study Overview

The aim of this study was to evaluate the social and economic impact of Misión Peniel’s cash transfer program, specifically with regards to understanding how this money was spent and whether it was enough to support recipients during a two-week quarantine. From July 2021 to mid-August 2021, we surveyed residents in Immokalee who had received a direct cash transfer between December 2020 to July 2021 from the local social support program developed by Misión Peniel. We conducted a structured interview where we asked a predefined list of questions, presented in the same order, with most questions being close-ended in nature with multiple options to choose from, and two open-ended follow-up questions to provide flexibility for answers that might not be covered by the pre-coded choices. The survey included questions on demographics (age, gender, race and ethnicity, marital status, education), employment (employment status, industry), food insecurity, housing insecurity (eviction notice, behind on rent or mortgage payment), transportation insecurity, income, financially supporting others, what the check was used for, whether the money was enough to meet 2 weeks’ financial needs, whether the money helped with quarantine in response to positive COVID-19 testing, whether the respondent quarantined for the recommended 10 days, whether the respondent required hospitalization, whether there were any difficulties and/or fees for cashing the check, preference for lump-sum distribution versus spaced distributions, previous experiences with cash transfer programs, etc. For respondents who responded “Other” to any given questions, we asked them to specify with details.

Study participants were contacted first by phone to explain the study and ask if they would like to participate. If we were not able to contact individuals by phone after two attempts, we also attempted two visits to the home address provided by Misión Peniel. If both phone and home visit contact were unsuccessful, we removed these individuals from the study. If someone could not be reached or chose not to participate, we pulled another individual from the list. Once contacted and verbally consented participants were read the survey questions either via phone or in person at their homes. The survey was administered in either English, Spanish, or Haitian Creole, depending on the respondent’s stated preference. Answers were entered into RedCap by the study investigators. This protocol was submitted to the University of Miami IRB office and deemed Not Human Research (IRB Submission 20210510).

### Eligibility Criteria

Participants who had been enrolled in Misión Peniel’s social support program and received a direct cash transfer were eligible for study participation. Only individuals ages 18 and older were included in this study. Individuals were excluded if they could not be contacted by either phone call or home visit. Misión Peniel provided a list of social support program recipients and their contact information. Each participant from the database was assigned a number, and a random number generator was used to select 275 individuals from this list out of 778 potential participants. This number was chosen by the researchers based on an anticipation of the maximum number of surveys that could be completed within the time frame of the study.

The final study sample consisted of 153 randomly selected Immokalee adult residents who either tested positive for COVID-19 and/or had a child living with them who tested positive for COVID-19, and who received a direct cash transfer from Misión Peniel during the time period of December 2020 to July 2021. The analysis sample is restricted to complete cases.

### Study Variables

#### Outcome Variable

The primary outcome variable was a dichotomous measure of whether the respondent reported the money was enough to cover their financial needs for two weeks.

#### Independent Variables

##### Food insecurity

During the interview, the respondents were asked whether they had experienced food insecurity; either not knowing where the next meal was coming from, or involuntarily eating less than they need, on a regular basis, for a period of time lasting more than a month) in the past 12 months.

##### Housing insecurity

During the interview, the respondents were asked two questions regarding housing insecurity. The first question was whether they received an eviction notice in the past 12 months; the second question was whether they had been behind on paying for rent or mortgage in the past 12 months.

##### Transportation insecurity

Respondents were also asked whether they experienced difficulty getting needed services because they did not have transportation in the past 12 months.

##### Income group

Respondents were asked about their average weekly income range, classified into five ordered categories including: 1) Less than $250; 2) $250-$400; 3) $400-$550; 4) $550-$700; and 5) More than $700. We created 3 income categories according to whether the converted annual income was below the 2021 national poverty line of $12,880 (approximately $12,900) for a household of one, between the poverty line and 2.2 times of the poverty line, or whether it exceeded 2.2 times of the poverty line. The value 2.2 is used here because the cut-off line between the third and fourth income category, which is a weekly income of $550, when converted into annual income, equals $12880, which is 2.2 times the federal poverty line. *Employment status*. Self-identified employment status was measured including 1) Employed (including full-time and part-time) or 2) Not working for pay/Unemployed.

### Covariates

A set of covariates including whether the check was used for purposes other than necessitates, whether the respondent was family caregiver, and demographic factors including gender, age, ethnicity, marital status, and educational level were controlled for in the model to sort out confounding relationships.

### Modeling Strategy

We fit a logit regression model on the data, in which food insecurity, housing insecurity, transportation insecurity, employment status, and income group are used to predict whether the money was enough for two weeks’ financial needs. We also include use of check, whether the respondent was currently financially supporting other individuals, and demographic indicators including gender, age, marital status, educational level, and race and ethnicity.

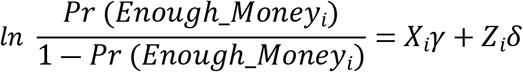

In the equation, *Enough*_*Money*_*i*_ indicates the binary outcome of whether the respondent thought the money was enough for two weeks’ financial needs. 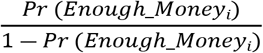 denotes the odds of the money being enough. 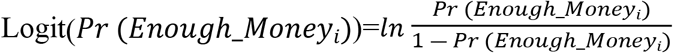 is the link function to convert the odds to the 0-1 interval. *X*_*i*_ is the matrix of predictors of respondent i; *Z*_*i*_ is the matrix of covariates of respondent i; *γ* and *δ* are vectors of regression coefficients. Subscript *i* indexes each individual observation.

## Results

A total of 157 Immokalee adult residents were surveyed out of 275 residents contacted (response rate of 57.1%). Of those surveyed, four respondents were excluded from the analysis due to missing income data. Table 1 represents all participant characteristics as well as two subgroups, created according to whether or not the money received met financial needs.

**Table 1.** Participant Characteristics.

| Variable | Full sample (N=153) | Whether the money was enough for 2 weeks |  |
| --- | --- | --- | --- |
|  |  | Yes (N=117) | No (N=36) |
| Age, mean [SD], years | 41 [14] | 41 [14] | 43 [13] |
| Male sex, N (%) | 97 (63.4) | 76 (65.0) | 21 (58.3) |
| Ethnicity, N (%) |  |  |  |
| Hispanic | 141 (92.2) | 108 (92.3) | 33 (91.7) |
| Non-Hispanic | 12 (7.8) | 9 (7.7) | 3 (8.3) |
| Married or cohabitated, N (%) | 73 (47.7) | 54 (46.2) | 19 (52.8) |
| Educational level, N (%) |  |  |  |
| Middle school or below | 75 (49.0) | 51 (43.6)** | 24 (66.7)** |
| High school or above | 78 (51.0) | 66 (56.4)** | 12 (33.3)** |
| Employment status, N (%) |  |  |  |
| Employed | 100 (65.4) | 74 (63.2) | 26 (72.2) |
| Not working for pay/unemployed | 53 (34.6) | 43 (36.8) | 10 (27.8) |
| Income group, N (%) |  |  |  |
| Income ≤ poverty line | 42 (27.5) | 33 (28.2) | 9 (25.0) |
| Income ≤ 2.2*poverty line | 81 (52.9) | 60 (51.3) | 21 (58.3) |
| Income $>$ 2.2*poverty line | 30 (19.6) | 24 (20.5) | 6 (16.7) |
| Currently financially supporting other individuals, N (%) | 108 (70.6) | 79 (67.5) | 29 (80.6) |
| Food insecurity, N (%) | 26 (17.0) | 15 (12.8)** | 11 (30.6)** |
| Housing insecurity, N (%) | 48 (31.4) | 29 (24.8)*** | 19 (52.8)*** |
| Transportation insecurity, N (%) | 26 (17.0) | 16 (13.7)** | 10 (27.8)** |
| Check use for purposes other than necessities, N (%) | 25 (16.3) | 22 (18.8) | 3 (8.3) |

The results of Chi-square tests for the differences in means of each variable for respondents who reported the money was or was not enough for two weeks' financial needs are denoted as asterisks next to the summary statistics.
\*\*\* $p < 0.01$ , \*\* $p < 0.05$ , \* $p < 0.1$ .

A majority of respondents (83.7%) reported spending the check exclusively on living expenses, including utilities (50.3%), food (45.9%), housing (44.6%), transportation (10.2%), and healthcare (8.9%). Of the recipients surveyed, 98.7% reported quarantining for ten days after their positive COVID-19 test, and 99.3% reported that the money helped them stay home and quarantine while having COVID-19. A total of 76.5% of respondents reported that the check was enough to cover their financial needs for two weeks. Table 2 presents the results from the logistic regression model in which employment status, income group, food insecurity, housing insecurity, and transportation insecurity are used as predictors of whether the respondent thought the money was enough for two weeks’ financial needs.

**Table 2.** Odds ratios of participant characteristics and whether the check was enough for 2 weeks’ financial needs.

| Variables | Odds Ratio (95% CI) |
| --- | --- |
| Age | 1.01 (0.97, 1.05) |
| Male | 0.78 (0.32, 1.86) |
| Hispanic | 1.11 (0.22, 5.68) |
| Married or cohabitated | 0.79 (0.29, 2.15) |
| High school education or above | 2.98** (1.03, 8.63) |
| Employed | 0.43 (0.13, 1.47) |
| Income group |  |
| Poverty line<Income≤2.2*Poverty line | 1.24 (0.32, 4.79) |
| Income>2.2*Poverty line | 1.12 (0.24, 5.31) |
| Experienced food insecurity in past 12 months | 0.42 (0.13, 1.34) |
| Experienced housing insecurity in past 12 months | 0.35** (0.15, 0.81) |
| Experienced transportation insecurity in past 12 months | 0.80 (0.24, 2.67) |
| Used check for purposes other than living necessities | 2.40 (0.65, 8.85) |
| Currently financially supporting individuals other than themselves | 0.88 (0.29, 2.72) |

Note: Estimates displayed in the form of odds ratios.
Confidence intervals are provided in parentheses.
\*\*\* p<0.01, \*\* p<0.05, \* p<0.1

Housing insecurity is negatively associated with the odds of reporting the money was enough for two weeks’ financial needs. Respondents who experienced housing insecurity in the past 12 months had a 65% decreased odds of reporting that the money was enough versus for respondents who did not experience housing insecurity. There was a significantly positive correlation between having a high school degree or above and the odds of reporting the money was enough. Respondents with a high school degree or above are 1.98 times more likely than respondents with middle school degree or less to report that the money was enough.

## Data Availability

All relevant data are within the manuscript and its Supporting Information files.

## Discussion

A common concern of policymakers with regards to cash transfers is that poor households will misuse the cash. This may explain why many governments and non-profit agencies opt for in-kind social assistance despite economic reasoning which suggests that cash transfers may be more efficient for beneficiaries [6]. This concern, however, is not supported by our data, wherein a vast majority of respondents (83.7%) reported spending the check exclusively on living necessities such as utilities, food, housing, transportation, and healthcare. These findings are supported by a literature review by Evans and Popova [7], which found a negative relationship between cash transfers and expenditures on temptation goods. Similar findings were seen also in another study examining one-time cash transfers of $1000 to individuals who tested positive for COVID-19 in New York from May 2020 to May 2021. The authors concluded that after receiving a cash transfer, individuals made “rational decisions to support their health and well-being rather than “misusing” funds on temptation goods such as alcohol” [8]. Additionally, our study revealed that nearly all recipients surveyed were able to self-isolate for 10 days as advised, which suggests that individuals in our sample had an awareness of and desire to comply with public health measures within their community.

Another measure in this study which may inform future programs was evaluating the quantity of cash support provided, which ranged from $800 to $1200. A majority of respondents (76.5%) reported that this amount was enough to cover their financial needs for two weeks. However, when examining these responses stratified by demographic variables, we found that people with housing insecurity and people without a high school degree were significantly less likely to report that the money was enough for two weeks’ financial needs (p<0.05). Therefore, these characteristics may mark those in the population who are especially vulnerable and could have benefitted from more support. One explanation is that housing insecurity may be one of the primary risk factors that prevents people from being able to self-isolate and stop spreading COVID-19 after getting a positive test. Some cash transfer programs have previously attempted to address disparities among recipients by creating targeted programs with varying eligibility requirements [9]. However, other studies have revealed that residency-based cash transfer programs may be more effective in practice than targeted programs, particularly during a pandemic, because they can act quickly without requiring the time and resources to verify who qualifies [10]. Therefore, we conclude that another way future cash transfer programs may increase the likelihood of adequately supporting vulnerable members of a population is by increased generosity in transfer amount.

One additional note is that a high number (81.7%) of respondents in our study answered “other” for the survey question regarding race. Race and ethnicity questions in this survey were based on the U.S. Census Bureau categories. This suggests that current census options exclude significant identities and may not be accurately capturing the race-ethnic composition of respondents, which has many implications for research utilizing these demographic measures.

## Limitations

One important limitation is that responses to questions about self-isolating and benefitting from the cash transfer may have been influenced by the social desirability bias, in which respondents may tend to offer answers that they believe will be viewed desirably by others rather than responses that are reflective of their true feelings or behaviors [11]. This is a significant concern, given that participants may have associated surveyors with those who provided the cash transfer. To mitigate the impact of this bias, it was explained prior to each survey that participant answers would not affect eligibility for future cash transfers, and that one aim of the survey was to identify areas of improvement within the program. Future studies should seek to evaluate these measures in comparison with a control group who did not receive funding. An additional limitation was the low response rate (57.1%). Many individuals were unable to be contacted even after two phone calls and two home visits, which may have been influenced in part by seasonal migration for work, as this study was conducted during the off-season for tomatoes in Immokalee. Our hypothesis is that this introduced a bias that over-sampled people who had fewer employment opportunities.

## Conclusion

As the COVID-19 pandemic continues to evolve it is essential to utilize best practices and policies to strategically address gaps in our public health response. It is indisputable that the pandemic has had a disproportionate impact on vulnerable and marginalized groups, particularly racial and ethnic minorities, and has exacerbated pre-existing disparities in the social determinants of health. Our study revealed that offering direct cash transfers of $800-$1200 to residents of Immokalee, FL who tested positive for COVID-19 was effective in reducing COVID-associated financial burden and demonstrated that money was most likely to be spent on living necessities rather than temptation goods. Therefore, we conclude that cash transfers may be one effective tool to strengthen community COVID-19 mitigation efforts and address health disparities and inequities.

## Supporting information

**S1 Table. Raw survey data**.

